# Community prevalence of SARS-CoV-2 in England during April to September 2020: Results from the ONS Coronavirus Infection Survey

**DOI:** 10.1101/2020.10.26.20219428

**Authors:** Koen B Pouwels, Thomas House, Emma Pritchard, Julie V Robotham, Paul J Birrell, Andrew Gelman, Karina-Doris Vihta, Nikola Bowers, Ian Boreham, Heledd Thomas, James Lewis, Iain Bell, John I Bell, John N Newton, Jeremy Farrar, Ian Diamond, Pete Benton, Ann Sarah Walker, COVID-19 Infection Survey team

## Abstract

**Background:** Decisions regarding the continued need for control measures to contain the spread of SARS-CoV-2 rely on accurate and up-to-date information about the number of people and risk factors for testing positive. Existing surveillance systems are not based on population samples and are generally not longitudinal in design.

**Methods:** From 26 April to 19 September2020, 514,794 samples from 123,497 individuals were collected from individuals aged 2 years and over from a representative sample of private households from England. Participants completed a questionnaire and nose and throat swab were taken. The percentage of individuals testing positive for SARS-CoV-2 RNA was estimated over time using dynamic multilevel regression and post-stratification, to account for potential residual non-representativeness. Potential changes in risk factors for testing positive over time were also evaluated using multilevel regression models.

**Findings:** Between 26 April and 19 September 2020, in total, results were available from 514,794 samples from 123,497 individuals, of which 489 were positive overall from 398 individuals. The percentage of people testing positive for SARS-CoV-2 changed substantially over time, with an initial decrease between end of April and June, followed by low levels during the summer, before marked increases end of August and September 2020. Having a patient-facing role and working outside your home were important risk factors for testing positive in the first period but not (yet) in the second period of increased positivity rates, and age (young adults) being an important driver of the second period of increased positivity rates. A substantial proportion of infections were in individuals not reporting symptoms (53%-70%, dependent on calendar time).

**Interpretation:** Important risk factors for testing positive varied substantially between the initial and second periods of higher positivity rates, and a substantial proportion of infections were in individuals not reporting symptoms, indicating that continued monitoring for SARS-CoV-2 in the community will be important for managing the epidemic moving forwards.

**Funding:** This study is funded by the Department of Health and Social Care. KBP, ASW, EP and JVR are supported by the National Institute for Health Research Health Protection Research Unit (NIHR HPRU) in Healthcare Associated Infections and Antimicrobial Resistance at the University of Oxford in partnership with Public Health England (PHE) (NIHR200915). AG is supported by U.S. National Institute of Health and Office of Naval Research. ASW is also supported by the NIHR Oxford Biomedical Research Centre and by core support from the Medical Research Council UK to the MRC Clinical Trials Unit [MC_UU_12023/22] and is an NIHR Senior Investigator. The views expressed are those of the authors and not necessarily those of the National Health Service, NIHR, Department of Health, or PHE.

**Research in context:** *Evidence before this study:* Unprecedented control measures, such as national lockdowns, have been widely implemented to contain the spread of SARS-CoV-2. Decisions regarding the continued need for social distancing measures in the overall population, specific subgroups and geographic areas heavily rely on accurate and up-to-date information about the number of people and risk factors for testing positive. We searched PubMed and medRxiv and bioRxiv preprint servers up to 6 June 2020 for epidemiological studies using the terms “SARS-CoV-2” and “prevalence” or “incidence” without data or language restrictions. Most studies were small or had only information about current presence of the virus for a small subset of patients, or used data not representative of the community, such as hospital admissions, deaths or self-reported symptoms. Large population-based studies, such as the current study, are required to understand risk factors and the dynamics of the epidemic.

*Added value of this study:* This is the first longitudinal community survey of SARS-CoV-2 infection at national and regional levels in the UK. With more than 500,000 swabs from more than 120,000 individuals this study provides robust evidence that the percentage of individuals from the general community in England testing positive for SARS-CoV-2 clearly declined between end of April and June 2020,, followed by consistently low levels during the summer, before marked increases end of August and September 2020. Risk factors for testing positive varied substantially between the initial and second periods of higher positivity rates, with having a patient-facing role and working outside your home being important risk factors in the first period but not (yet) in the second period, and age (young adults) being an important driver of the second period of increased positivity rates. Positive tests commonly occurred without symptoms being reported.

*Implications of all the available evidence:* The observed decline in the percentage of individuals testing positive adds to the increasing body of empirical evidence and theoretical models that suggest that the lockdown imposed on 23 March 2020 in England was associated, at least temporarily, with a decrease in infections. Important risk factors for testing positive varied substantially between the initial and second periods of higher positivity rates, and a substantial proportion of infections were in individuals not reporting symptoms, indicating that continued monitoring for SARS-CoV-2 in the community will be important for managing the epidemic moving forwards.

## Introduction

Since severe acute respiratory syndrome coronavirus 2 (SARS-CoV-2) started causing severe respiratory illness in Wuhan, China, in late 2019,^1^ the virus has had a drastic impact worldwide. As of 20 September, there have been over 30.6 million confirmed cases and 950,000 deaths reported to the WHO.^2^ Control measures, such as national lockdowns, have been widely implemented to contain the spread of the virus in a, at least temporarily successful,^3-5^ attempt to prevent the collapse of healthcare systems and even larger numbers of deaths among those infected with SARS-CoV-2. Although such measures are important for control of the pandemic, they also affect the economy, unemployment rates, and global supply-chains.^6,7^ Politicians continuously make the difficult decision between continuing strict control measures or relaxing them in some way that would be safe enough from a public health perspective yet beneficial more broadly across society.

Importantly, early detection of population subgroups driving new increases in infections is crucial to decisions around potential tailoring interventions or messaging without having to implement drastic measures affecting the whole society.

There are several reasons why risk factors may vary over time. First, behaviour and contact patterns of subgroups change over time without intervention, e.g. students starting university. Adherence to non-mandatory infection prevention measures may reduce more over time among subgroups with a low risk of COVID-19 related hospital admission and death than those that are more vulnerable. Moreover, subgroups that have been disproportionally affected in a first wave may have acquired sufficient immunity and may have better access to effective measures that reduce the risk of infection making them less likely to acquire a new infection during a second wave.

Here, we use data from the Office of National Statistics (ONS) Coronavirus Infection Survey (CIS). This large national survey with more than 500,000 swab results to 19 September is designed to be representative of the target population, offering a unique opportunity to identify risk factors that are driving recent new increases in the positivity rate, as well as investigating the proportion of individuals testing positive that do not report symptoms, potential false-positivity rate, and other factors that can directly inform policy around COVID-19 related control measures. We used Bayesian dynamic multilevel regression and poststratification (MRP) to account for any residual unrepresentativeness, a potential problem often ignored with surveillance data.

## Methods

Data were collected between 26 April and 19 September 2020 from individuals from randomly selected private households from address lists and previous ONS surveys to provide a representative sample of the population of England. Only individuals aged 2 years and older living in private households were eligible for inclusion in the survey. If one or more individuals from a household agreed to participate, a visit by a study worker was arranged and individuals were asked about any symptoms and contacts, together with information about their gender, age, ethnicity and occupation. These questions were collected directly by the study worker. The study worker provided instructions on how to self-swab the nose and throat, which has been shown to be comparable or even more sensitive than swabs performed by healthcare workers.^9^ Parents/carers took swabs from children under 12 years old. The nose and throat self-swabs were couriered directly to the UK’s national Lighthouse laboratories at Milton Keynes (National Biocentre) (from 26 April) and Glasgow (from 16 August), where the samples were tested using identical methodology for the presence of SARS-CoV-2 (3 gene targets) using reverse transcriptase polymerase chain reaction (RT-PCR) as part of the national testing programme.^10^ After the first visit, participants were asked whether they were willing to participate in further follow-up visits: every week for the first 5 weeks of the study, and then monthly thereafter. The study protocol is available at https://www.ndm.ox.ac.uk/covid-19/covid-19-infection-survey/protocol-and-information-sheets. The project has been reviewed and given ethical approval by South Central - Berkshire B Research Ethics Committee (20/SC/0195).

### Trend in proportion of positive tests over time

We analysed the proportion of the private-residential population testing positive for SARS-CoV-2 from nose and throat swabs over time using Bayesian dynamic Multilevel Regression and Poststratification (MRP).^11,12^ MRP was used to correct for any residual non-representativeness in terms of age, sex and region. In several empirical and simulation studies MRP was be superior at both the national and regional levels compared to classical survey weighted and unweighted approaches, including when using small sample sizes.^11-16^ Partial pooling through the use of random effects in the multilevel model ensures stable estimates can be obtained for subnational levels from relatively small samples that would be problematic using more traditional survey-weighting approaches.^11-16^ MRP consists of two steps. First, a multilevel regression model is used to generate the outcome of interest as a function of (socio)demographic and geographic variables. Next, the resulting outcome estimates for each demographic-geographic respondent type are poststratified by the percentage of each type in the actual overall population.^11^

We used a Bayesian multilevel generalised additive regression model to model the swab test result (positive/negative) as a function of age, sex, time and region. Because there were very few missing values (≤1%) in these factors, we restricted all analyses to observations with non-missing data. A complementary log-log link was used due to the ability to interpret regression coefficients as arising from an infection process with varying levels of exposure (see Supplementary File).^17^ MRP models with random effects for individual participant and/or household nested within region did not converge. Therefore MRP models were run with only a random intercept for region, without a random intercept for participant and/or household. However, a model with only one participant sampled from each household gave similar results with somewhat wider 95% credible intervals mainly due to the smaller sample size (Figure S1). Time, measured in days since the start of the study (26 April 2020), was modelled using thin plate splines and allowed to vary by region. We set **k**, the number of basis functions, to to control the smoothness of the fitted function.^18^ We use a normal prior with location set to 4 for the standard deviation of the smooth. Very similar results were obtained when using different values for k (Figure S2A) or different priors for the standard deviation of the smooth (Figure S2B).Subsequently, we poststratified the resulting positivity estimates for each demographic-geographic respondent type by the percentage of each type in the overall population and in each region. This analysis was performed using the rstanarm package in R version 3.6.1.^19^

### Time-varying risk factors

To assess whether particular subgroups were more likely to test positive for SARS-CoV-2 viral RNA during the first wave in England we performed a multilevel regression analysis (without poststratification) on the data between 26 April and 28 June 2020 including variables on which we did not post-stratify: work location, having a job that directly involved patients/care-home residents, ethnicity, household size, and number of children in the household. Given the short timescale included, and the fact that questions were not always asked at every visits, we carried non-missing data forward and backwards to adjacent visits with missing data. After this, there were very few missing values (≤1%) so we again restricted all analyses to observations with non-missing data only. Results are shown in Table S1 and suggest that confounding plays a limited role.

During the summer positivity rate remained low and approximately constant, but more recently started to increase again. We evaluated using Bayesian dynamic multilevel regression models to what extent different factors were potentially driving this recent increase. Given that age appeared to be such a strong factor in driving the increase (see Results), all other factors were subsequently stratified by age (<35 and 35+ years). We evaluated the same factors as for the first wave (data up to 28 June) in generalised additive models with thin-plate splines that varied by each level of the factor of interest. These models additionally included a random intercept for region to account for any regional differences. As it was not possible to fit all factors with these time interactions in one model, and given the limited evidence of confounding (Table S1), we fitted separate models for each factor of interest.

### Presence of symptoms among those testing positive

We evaluated the number of positive tests where the participant reported that they had symptoms around the visit (same visit or visit before or after) or not using the same MRP model as for the overall positivity rate.

To assess the impact of potential false positive tests we classified each positive into 3 categories:

i. ‘Higher evidence’; two or three genes detected (irrespective of cycle threshold (Ct) value).
ii. ‘Moderate evidence’; single gene detections if a) the Ct value was <97.5^th^ percentile of ‘higher evidence’ positives (<34) or b) there was a higher pre-test probability of infection, i.e. any symptoms at or around the test (visit before or after) or reporting working in patient-facing healthcare role or resident-facing care home role.
iii. ‘Lower evidence’; all other positives, which by definition were all in asymptomatic individuals not having patient- or resident-facing roles with a single gene detected with Ct ≥34.

## Results

Between 26 April and 19 September 2020, in total, results were available from 514,794 samples from 123,497 individuals, of which 489 were positive overall from 398 individuals in 342 households. The sample was broadly representative in terms of age, sex, and region (Figure S1). Small under/overrepresentation of certain groups, such as individuals from London being slightly underrepresented, was corrected for using dynamic MRP with poststratification performed on a daily basis. Positivity rates dropped to consistently low levels during the summer before increasing markedly again end of August and September (Figure 1).

**Figure 1.**
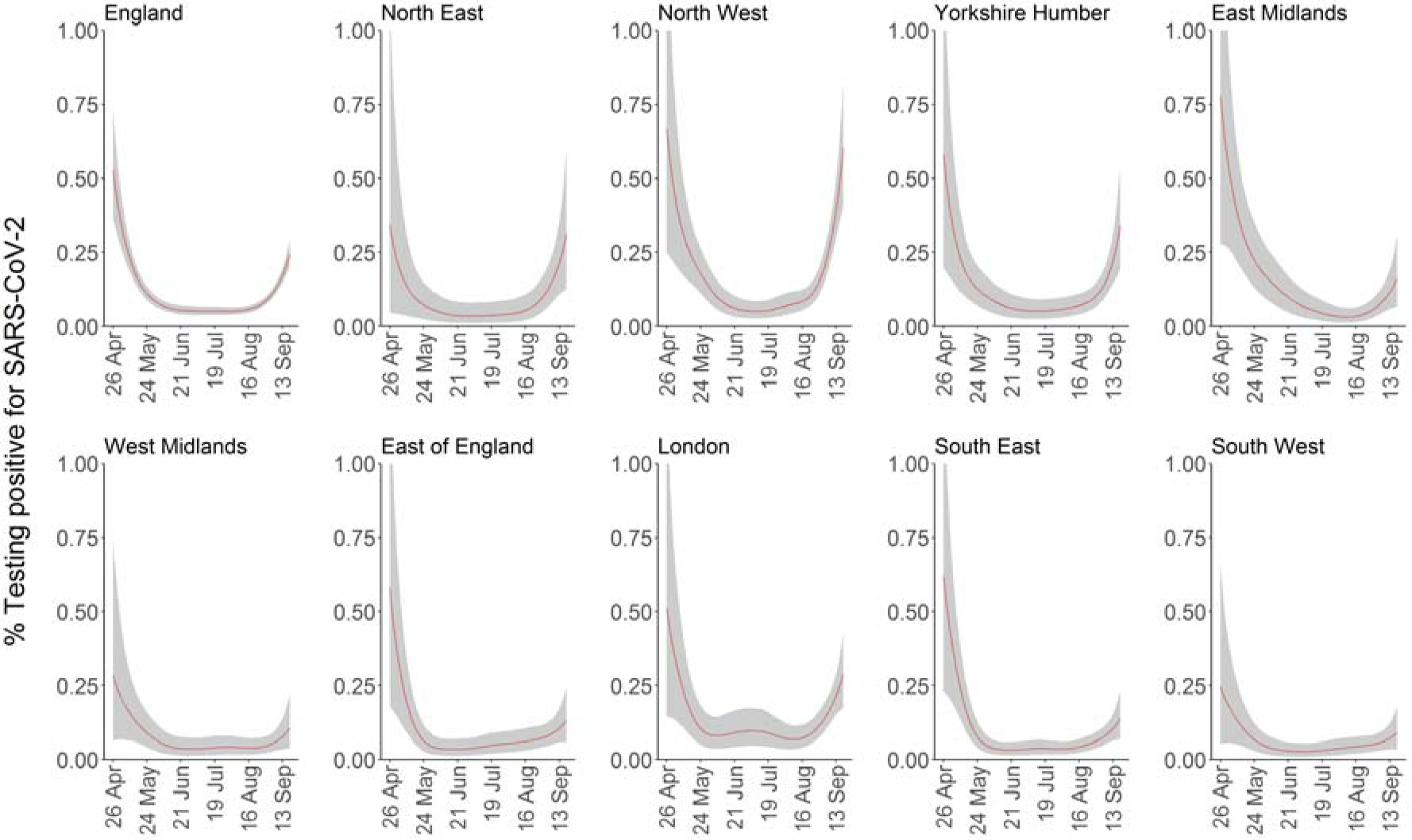
Percentage of population living in private households testing positive for SARS-CoV-2 over time in England and the 9 regions of England. Shaded areas are 95% credible intervals.

Similar patterns were observed for the positivity rate when the participant reported symptoms or not (Figure 2A. In July there was a slight temporary increase in the positivity rate without symptoms, while the positivity rate with symptoms remained approximately constant. The modelled percentage of positives with reported symptoms around the test was lowest around mid-July with 30% and highest around the end of the study period at 46%. The rate of lower evidence positives remained approximate stable over the study period, although again with a small increase during the summer, while the sharp increase around September 2020 was almost entirely due to an increase in higher evidence positives (Figure 2B), suggesting that people may have become infected with lower viral loads and fewer symptoms during the summer, but with higher viral loads in September potentially leading to a higher proportion of cases with symptoms.

**Figure 2.**
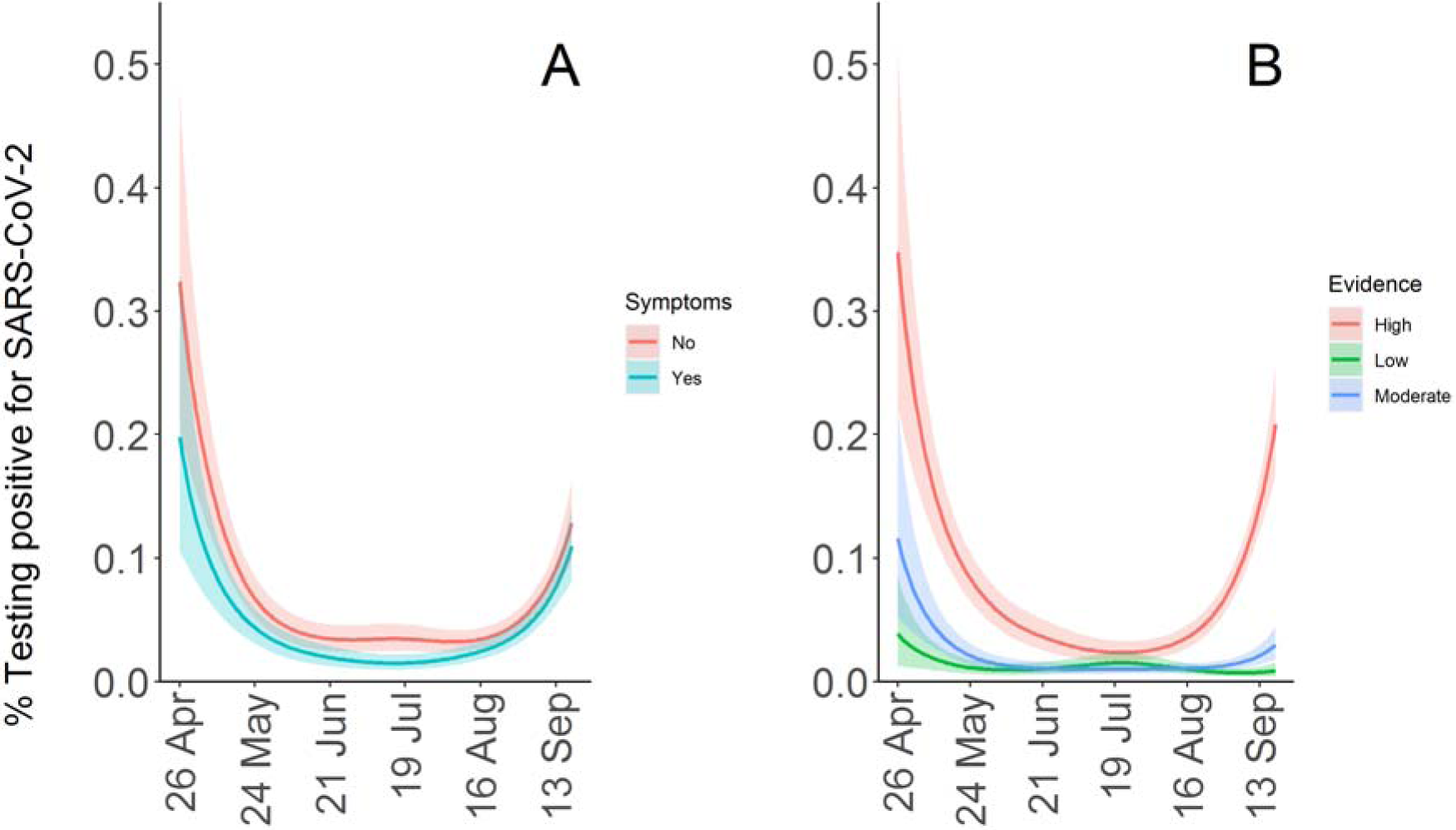
**A:** Percentage of population living in private households testing positive for SARS-CoV-2 with and without reporting symptoms; **B**:Percentage of population living in private households testing positive for SARS-CoV-2 stratified by high, moderate and low evidence positivity. Shaded areas are 95% credible intervals.

Positivity rates showed marked regional differences, with increases in late August-September largely occurring in the North of England (Figure 1). We explored whether other factors besides region may be underlying the observed sharp increase in September 2020 using dynamic generalised additive mixed models without poststratification since population distributions were unknown for most of these factors. The most important factor was age, with earlier and greater increases apparent in younger adults (Figure 3) as also evident from results from a model categorising age (Figure S4). Importantly, there was clear diffusion of risk from initial increases in younger age groups at lower risk of hospitalisation and death, into older ages at higher risk.

**Figure 3.**
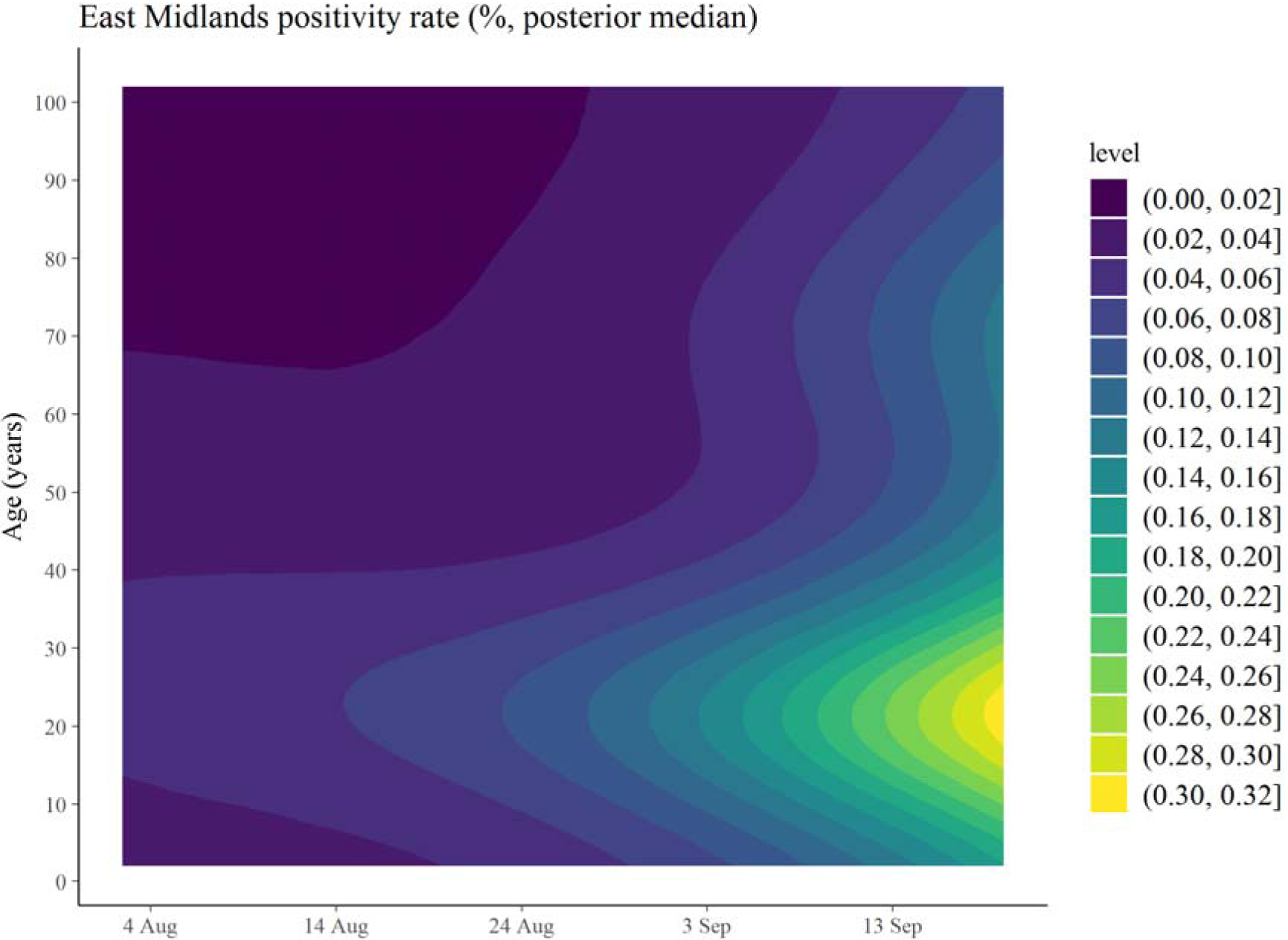
Modelled estimates (posterior medians) of the distribution of positive SARS-CoV-2 tests across age over time.

While working outside your home and in patient-facing healthcare roles were a clear risk factor during the initial period of high positivity, as was contact with hospitals (26 April to 28 June, Table S1), there was no evidence that those working outside their home, working in patient-facing roles or with hospital contact were driving increases after the summer (Figure S5-7). Whilst non-White ethnicity was also associated with greater positivity rates during the initial period (Figure S8), increases after the summer were greatest in White individuals under 35 years (Figure 4). In contrast, there were different trends in the most recent period dependent on household composition, where increases were more apparent in households with children of both primary and secondary school age and households consisting of one person age between 17 and 34 than in other categories (Figure 5).

**Figure 4.**
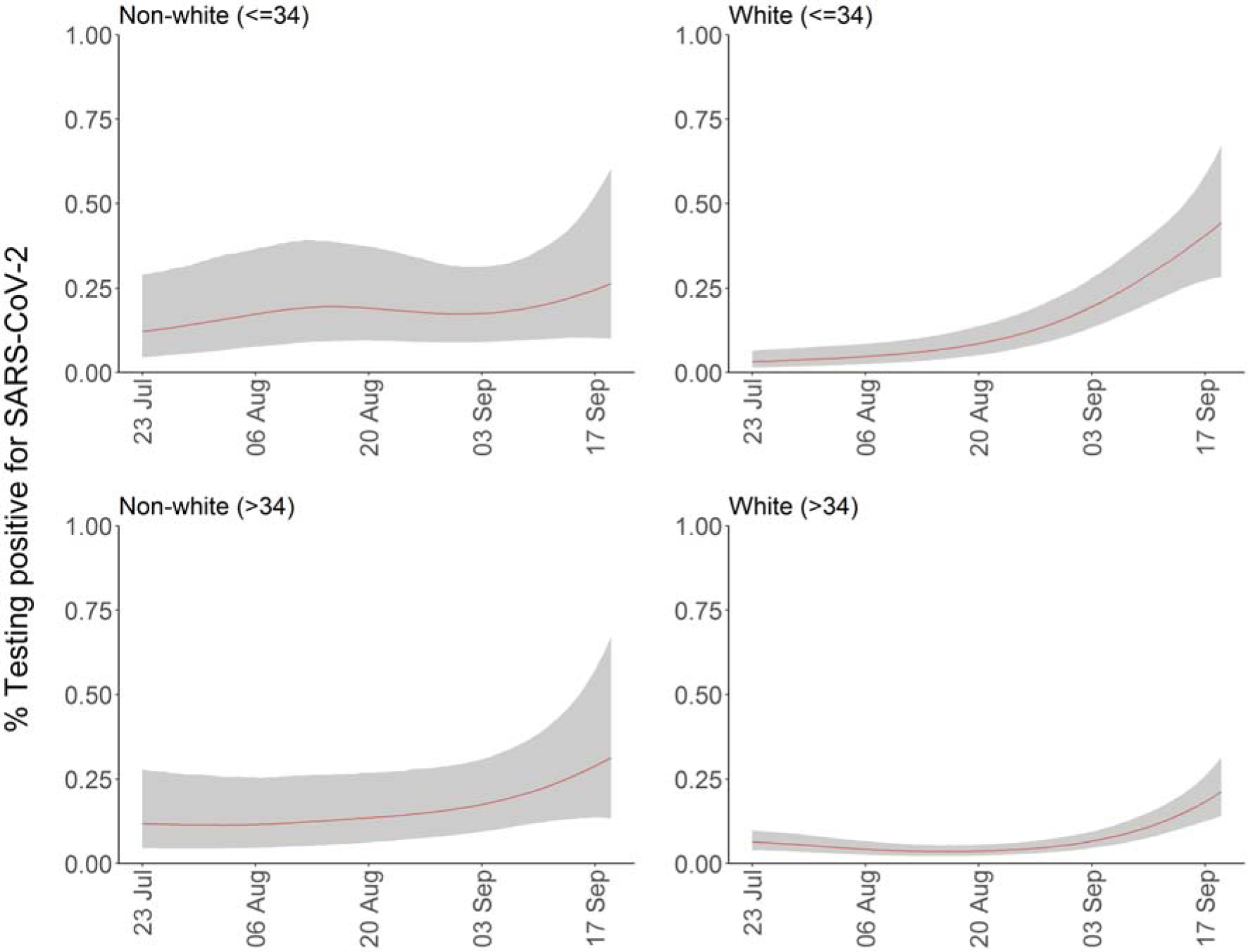
Percentage of population living in private households testing positive for SARS-CoV-2 stratified by ethnicity and age (<=34 and >34 years of age). Shaded areas are 95% credible intervals.

**Figure 5.**
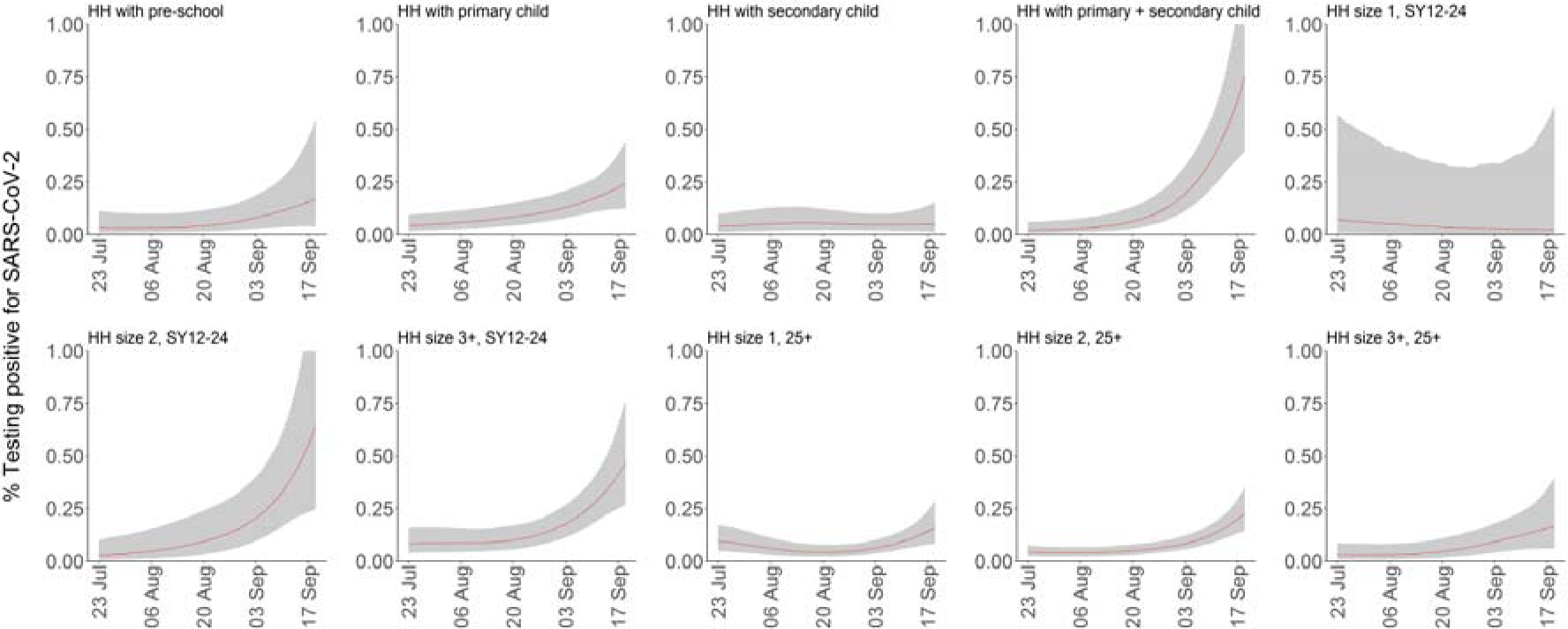
Percentage of population living in private households testing positive for SARS-CoV-2 stratified by their household composition. HH=household; SY=school year; SY12-24 indicates individuals from school-year 12 (aged 16/17) to age of 24 years old. Shaded areas are 95% credible intervals.

## Discussion

Here we demonstrate substantial changes over time in the percentage of people in private-residential households in the community in England testing positive for SARS-CoV-2, with an initial decrease between end of April and June 2020, followed by consistently low levels during the summer, before marked increases end of August and September 2020. Our estimates have been regularly updated and shared with the UK Government and Scientific Advisory Group for Emergencies (SAGE) sub-group Scientific Pandemic Influenza sub-group on Modelling (SPI-M) to directly inform decisions about potential changes to the current alert level or relaxation of certain restrictions.

Notably, we found that a substantial proportion (53%-70%, dependent on calendar time) of individuals that tested positive did not report any symptoms on the day of the visit or at visits before or after the swab was taken.

The Bayesian dynamic multilevel generalised additive models are useful tools for monitoring the effect of different factors on positivity rates over time. In particular, they show that the epidemic restarted in young people, and that factors associated with an increased risk of testing positive during the initial high-prevalence period in April-May 2020, such as working outside the home and having a job with direct patient contact were not important drivers of the recent increase after the summer.

While false-positives may be a concern when prevalence is low, the low prevalence at the end of June (0.05%) is also reassuring, since it indicates that the specificity of the test used in the national UK programme is very high. A test specificity lower than 99.95% would lead to observed positivity rates above 0.05%, even in the purely hypothetical situation that the virus was not circulating in June. In theory, the percentage of tests that are false-positives might be expected to be relatively stable over time. Interestingly, the number of lower-evidence positives, i.e. with a low pre-test probability and a single gene detected with Ct value ≥34, and hence more likely to be false positives, increased slightly during the summer. This may be partly a consequence of a genuinely increase in the number of infections with low levels of virus during this period.

### Comparison with other studies

Our finding of a temporal reduction in the percentage of people in the community infected with SARS-CoV-2 in England between April and June 2020 is consistent with reductions observed in the weekly number of excess deaths and COVID-19 hospitalisations.^20,21^ An important advantage of our current population-based study is that it can detect increases in the positivity rate potentially earlier and more systematically than surveillance based on confirmed cases, hospital admissions or deaths (https://coronavirus.data.gov.uk/). This is likely especially the case when new increases initially occur in a subgroup of the population that is at low risk of hospitalisation and death, but does contribute to transmission including if asymptomatic,^22^ as observed after the summer with the increase in positives among young adults. Furthermore, interpretation of changes in incidence and positivity rate from tests that are taken for contact tracing or clinical cases is likely confounded by substantial changes in testing practice over time. Our study is based on a representative sample of the population, with further correction for residual non-representativeness using MRP, thereby preventing difficulties with interpretation due to changes in testing practice.

There are a few other studies that aimed to assess the prevalence of SARS-CoV-2 infection in the general population. A repeated cross-sectional population-based study from England also found a similar decline in the prevalence among the general population between 1 May and 1 June.^23^ Another cross-section from that study showed also an increase in in the prevalence in September.^24^ Among individuals that tested positive in that study, the percentage reporting no symptoms varied between 50% and 81% between the different cross-sections.^24^ A study from Vo, an Italian town with a population of 3275 individuals, Lavezzo et al. surveyed 85.9% and 71.5% of the inhabitants at the start and end of lockdown of the town respectively, with an initial infection prevalence of 2.6% (95% confidence interval (CI) 2.1-3.3%) and 1.2% (95% CI 0.8-1.8%) 14 days later.^25^ The percentage of those who tested positive that did not report any symptoms was 41.0% (95% CI 29.7-53.2%) in the first survey and 44.8% (95% CI 26.5-64.3%) in the second survey.

As part of a larger study from Iceland, a randomly selected sample of inhabitants aged between 20 and 70 years of age were invited to participate in a survey.^26^ By 4 April, 2283 (33.7%) of those invited had participated, with 13 testing positive (0.6%; 95% CI 0.3%-1.0%). However, it is unclear to what extent the participation rate led to a sample that was not representative of the Icelandic population and no correction for population representativeness was performed. Among a larger sample, including participants recruited via an open invitation, which may bias the sample towards people with symptoms, 57% of individuals testing positive reported having symptoms, although 29% of individuals testing negative also reported having symptoms.

While the studies from Iceland and Vo found that around 40-45% of those with a positive test did not report any symptoms,^25,26^ this percentage is higher in our study. This may be partly due to differences in respondents, chance (given considerable uncertainty in all studies), differences in definitions of symptoms, over/under-reporting of symptoms or false-positive tests.

Furthermore, the similar trends over time for positives with and without symptoms suggests that there truly may be more asymptomatic cases than reported in some other studies.^27^ A recent meta-analysis of studies focusing on close contacts of confirmed COVID-19 cases suggested that only 17% (95% CI 14%-20%) of infected individuals are asymptomatic.^27^ However, by informing participants that they were recently in close contact with a confirmed COVID-19 case, individuals may be more likely to think that they have experienced symptoms, resulting in recall bias and overestimating the true prevalence of symptoms among a representative sample of infected persons. Although we may have underestimated the true prevalence of symptoms among SARS-CoV-2 cases in the community, partly due to asking about current symptoms at visits through 23 July (meaning that very transient symptoms only occurring between visits would have been missed) and symptoms in the last 7 days thereafter, our study adds to the growing evidence that a substantial proportion of SARS-CoV-2 in the community may be asymptomatic.^28,29^

### Limitations of this study

An important limitation of this study is that the number of people in the community that test positive is low, limiting power and leading to relatively large uncertainty around estimates, and meaning that our multilevel regression model was not able to incorporate likely correlation within households. However, sensitivity analyses suggested that within-household clustering did not have a large impact on our results.

Furthermore, while we adjusted for potential non-representativeness in terms of age, sex and region, there may be other factors for which we do not have detailed information about population distributions that also are associated with testing positive. If these are over-or under-represented in the survey this could have resulted in some residual bias. We did forwards and backwards imputation for missing data, reflecting the relatively short timescales of the study.

Another limitation is that, in the absence of a true gold standard, we do not know the test sensitivity and specificity, making it difficult to assess what the true prevalence is. However, as detailed above the true specificity is likely very close to 100%The data cannot inform about the test sensitivity without providing a very informative prior on the true prevalence.^30^ However, this should not affect trends in prevalence over time.

## Conclusions

The percentage of individuals from the community in England testing positive for SARS-CoV-2 clearly declined between 26 April and 28 June 2020, remained approximately stable for much of the summer before increasing again from the end of August through September. Important risk factors for testing positive varied substantially between the initial and second periods of higher positivity rates, and a substantial proportion of infections were in individuals not reporting symptoms, indicating that continued monitoring for SARS-CoV-2 in the community will be important for managing the epidemic moving forwards. Bayesian dynamic Multilevel Regression and Poststratification (MRP) is a powerful tool which could be used more widely to ensure population-representativeness of surveillance estimates.

## Data Availability

De-identified study data are available for access by accredited researchers in the ONS Secure Research Service (SRS) for accredited research purposes under part 5, chapter 5 of the Digital Economy Act 2017. For further information about accreditation, contact or visit the SRS website

https://www.ons.gov.uk/aboutus/whatwedo/statistics/requestingstatistics/approvedresearcherscheme

https://www.statisticsauthority.gov.uk/about-the-authority/better-useofdata-statistics-and-research/betterdataaccess-research/

## Contributors

The study was designed and planned by SW, JF, JB, JN, IB, ID, PB, KBP, and JVR. KBP, TH, EP, KDV, NB, IB, HT, JL and ASW contributed to the statistical analysis. KBP drafted the manuscript and all authors contributed to interpretation of the data and results and revised the manuscript. KBP and ASW are the guarantors of the study. All authors approved the final version of the manuscript and agreed to be accountable for all aspects of the work. The corresponding authors attests that all listed authors meet authorship criteria and that no others meeting the criteria have been omitted.

## Declaration of interest

We declare no competing interests.

## Data sharing

Data sharing: De-identified study data are available for access by accredited researchers in the ONS Secure Research Service (SRS) for <u>accredited research purposes</u> under part 5, chapter 5 of the Digital Economy Act 2017. For further information about accreditation, contact or visit the <u>SRS website</u>

## Acknowledgements

**COVID-19 Infection Survey Team**

**COVID-19 Infection Survey Team**

**Office for National Statistics:** Iain Bell, Ian Diamond, Alex Lambert, Pete Benton, Emma Rourke, Stacey Hawkes, Sarah Henry, James Scruton, Peter Stokes, Tina Thomas.

**Office for National Statistics, Analysis:** John Allen, Russell Black, Heather Bovill, David Braunholtz, Dominic Brown, Sarah Collyer, Megan Crees, Colin Daglish, Byron Davies, Hannah Donnarumma, Julia Douglas-Mann, Antonio Felton, Hannah Finselbach, Eleanor Fordham, Alberta Ipser, Joe Jenkins, Joel Jones, Katherine Kent, Geeta Kerai, Lina Lloyd, Victoria Masding, Ellie Osborn, Alpi Patel, Elizabeth Pereira, Tristan Pett, Melissa Randall, Donna Reeve, Palvi Shah, Ruth Snook, Ruth Studley, Esther Sutherland, Eliza Swinn, Heledd Thomas, Anna Tudor, Joshua Weston.

**Office for National Statistics, Secure Research Service:** Shayla Leib, James Tierney, Gabor Farkas, Raf Cobb, Folkert van Galen, Lewis Compton, James Irving, John Clarke, Rachel Mullis, Lorraine Ireland, Diana Airimitoaie, Charlotte Nash, Danielle Cox, Sarah Fisher, Zoe Moore, James McLean, Matt Kerby.

**University of Oxford, Nuffield Department of Medicine:** Ann Sarah Walker, Derrick Crook, Philippa C Matthews, Tim Peto, Emma Pritchard, Nicole Stoesser, Karina-Doris Vihta, Alison Howarth, George Doherty, James Kavanagh, Kevin K Chau, Stephanie B Hatch, Daniel Ebner, Lucas Martins Ferreira, Thomas Christott, Brian D Marsden, Wanwisa Dejnirattisai, Juthathip Mongkolsapaya, Sarah Hoosdally, Richard Cornall, David I Stuart, Gavin Screaton.

**University of Oxford, Nuffield Department of Population Health:** Koen Pouwels.

**University of Oxford, Big Data Institute:** David W Eyre.

**University of Oxford, Radcliffe Department of Medicine:** John Bell.

**Oxford University Hospitals NHS Foundation Trust:** Stuart Cox, Kevin Paddon, Tim James.

**University of Manchester:** Thomas House.

**Public Health England:** John Newton, Julie Robotham, Paul Birrell.

**IQVIA:** Helena Jordan, Tim Sheppard, Graham Athey, Dan Moody, Leigh Curry, Pamela Brereton

**National Biocentre:** Ian Jarvis, Kirsty Howell, Bobby Mallick, Phil Eeles.

**Glasgow Lighthouse Laboratory:** Jodie Hay, Harper Vansteenhouse.

## Notes

### Competing Interest Statement

The authors have declared no competing interest.

### Author Declarations

The project has been reviewed and given ethical approval by South Central - Berkshire B Research Ethics Committee (20/SC/0195).

