## Supplementary Files for "Community prevalence of SARS-CoV-2 in England during April to September 2020: Results from the ONS Coronavirus Infection Survey"

#### 1 Models estimated in the paper

*Dynamic MRP*

The regression model that was used for the dynamic multilevel model and post-stratification (MRP) analysis was a Bayesian multilevel generalised additive model (GAMM) with a complementary loglog link implemented using the rstanarm package.[1-2] Sex was modelled as a fixed effect as it has only 2 levels, while age (5 levels) and region (12 levels) were modelled as random effects. This model was implemented using the following syntax:

```
stan_gamm4(result ~ s(time, by=region, k=10) + sex,
random = ~(1| age) + (1|region),
family = binomial(link="cloglog"),
data = data, iter = 3000, cores = 4,
prior = normal(0,0.5), prior_covariance
= decov(shape = 1, scale = 1),
prior_smooth=normal(location=4),
control=list(adapt_delta=0.95))
```

*Associations between variables and testing positive*

To assess whether particular subgroups are more likely to test positive for SARS-CoV-2 viral RNA we performed an additional analysis including variables on which we did not post-stratify. We used the same model as for the dynamic MRP but in working aged individuals only (16-74 years inclusive as defined in the Labour Force Survey) with these additional variables included as fixed covariates and age modelled as a continuous variable, using a thin-plate spline, instead of a categorical variable. Associated results can be found in Table S1.

#### 2 Epidemiological interpretation of the complementary log-log link function when focusing on associations between variables and testing positive for the presence of SARS-CoV-2 RNA.

Our regression model operates at the individual level; in particular, we assume that there are $n$ swabs taken, and the $i$-th of these is associated with time $t_i$, English region $e_i$, and a vector of other covariates $x_i$. These covariates are as detailed in the main text: age; work etc. The probability of the $i$-th swab being positive is then given by a generalised linear model (generalised additive model).

$$\pi_i = p(x_i, e_i, t_i) = g^{-1}\left(s_{e_i}(t_i) + \beta \cdot x_i + \zeta_{e_i}\right). \quad (1)$$

Here, $g$ is the link function of the GAMM, $s_{e_i}$ is the time smoother for the region $e$, $\beta$ is the vector of regression coefficients, and $\zeta_e$ is the random effect for region $e$, with these effects assumed i.i.d. with $\zeta_e \sim N(0, \sigma^2)$. The likelihood function for this model given observations $y_i = 1$ for a positive swab and $y_i = 0$ for negative, is then

$$L = \prod_{i=1}^{n} \pi_i^{y_i}(1 - \pi_i)^{1-y_i}. \quad (2)$$

Now suppose that the individual $i$ acquires infection at a rate $\lambda_i(t)$, known as the *force of infection* in infectious disease modelling. The Chapman-Kolmogorov equation, using dots for time derivatives, is:

$$\dot{\pi}_i(t) = (1 - \pi_i)\lambda_i(t). \quad (3)$$

This has solution

$$\pi_i = 1 - \exp(-\int_{u=0}^{t} \lambda_i(u)du). \quad (4)$$

If we choose a complementary log-log link function

$$g(x) = \log(-\log(1 - x)) \quad (5)$$

in (1), and assume that

$$\lambda_i(t) = \emptyset_i \lambda_{e_i}(t). \quad (6)$$

in (5), then we get

$$\pi_i = 1 - \exp\,(-\emptyset \int_{u=0}^{t} \lambda_{e_i}(u)du) = 1 - \exp\left(-\,exp(\beta \cdot x_i)\,exp\left(s_{e_i}(t_i)\right)exp\left(\zeta_{e_i}\right)\right). \quad (7)$$

This implies that

$$\emptyset_i \propto \prod_a e^{\beta_a x_{ia}}, \tag{8}$$

and so we quote the value of exp($\beta_a$) for the $a$-th covariate, with 1 as reference, since this is interpretable as the relative exposure to infectious risk.

**Table S1.** Risk factors for testing positive for SARS-CoV-2 between 26 April and 28 June 2020.

| Factor | Number of visits in sample (number positive) | Relative exposure to SARS-CoV-2 (95% CrI)[a] |
|---|---|---|
| Male | 44,308 (53) | Ref. |
| Female | 49,308 (57) | 0.84 (0.57 - 1.25) |
| *Work location* | | |
| Working from home | 22,392 (11) | Ref. |
| Working outside of your home | 20,621 (45) | 2.47 (1.40 - 4.55) |
| Both | 4,370 (5) | 1.43 (0.53 - 3.54) |
| Not applicable | 46,233 (49) | 2.09 (1.20 - 3.75) |
| *Job with direct contact with patients or care home residents* | | |
| Non-patient facing | 89,643 (87) | Ref. |
| Patient facing | 3,973 (23) | 4.06 (2.37 - 6.72) |
| Non-resident facing | 92,690 (105) | Ref. |
| Care home resident facing | 926 (5) | 2.35 (0.85 - 5.27) |
| *Ethnicity* | | |
| White | 88,793 (93) | Ref. |
| Asian | 2,514 (8) | 1.89 (0.87 - 3.64) |
| Black | 788 (2) | 1.04 (0.28 - 3.07) |
| Mixed | 1,068 (0) | 0.46 (0.09 - 1.84) |
| Other | 453 (7) | 7.50 (2.86 - 16.50) |
| *Household size* | | |
| 1 | 13,096 (16) | Ref. |
| 2 | 40,426 (28) | 0.62 (0.35 - 1.09) |
| 3 | 17,125 (33) | 1.51 (0.83 - 2.73) |
| 4 | 16,704 (21) | 1.36 (0.68 - 2.63 |
| 5 or more | 6,261 (12) | 1.25 (0.51 - 2.88) |
| *Number of children in household* | | |
| 0 | 64,917 (70) | Ref. |
| 1 | 11,206 (22) | 1.01 (0.59 - 1.73) |
| 2 | 10,083 (8) | 0.44 (0.20 - 0.94) |
| 3 or more | 2,593 (8) | 1.65 (0.64 - 4.17) |

[a]A relative exposure of 1 is the reference value (no effect).

### 3 Figures and tables

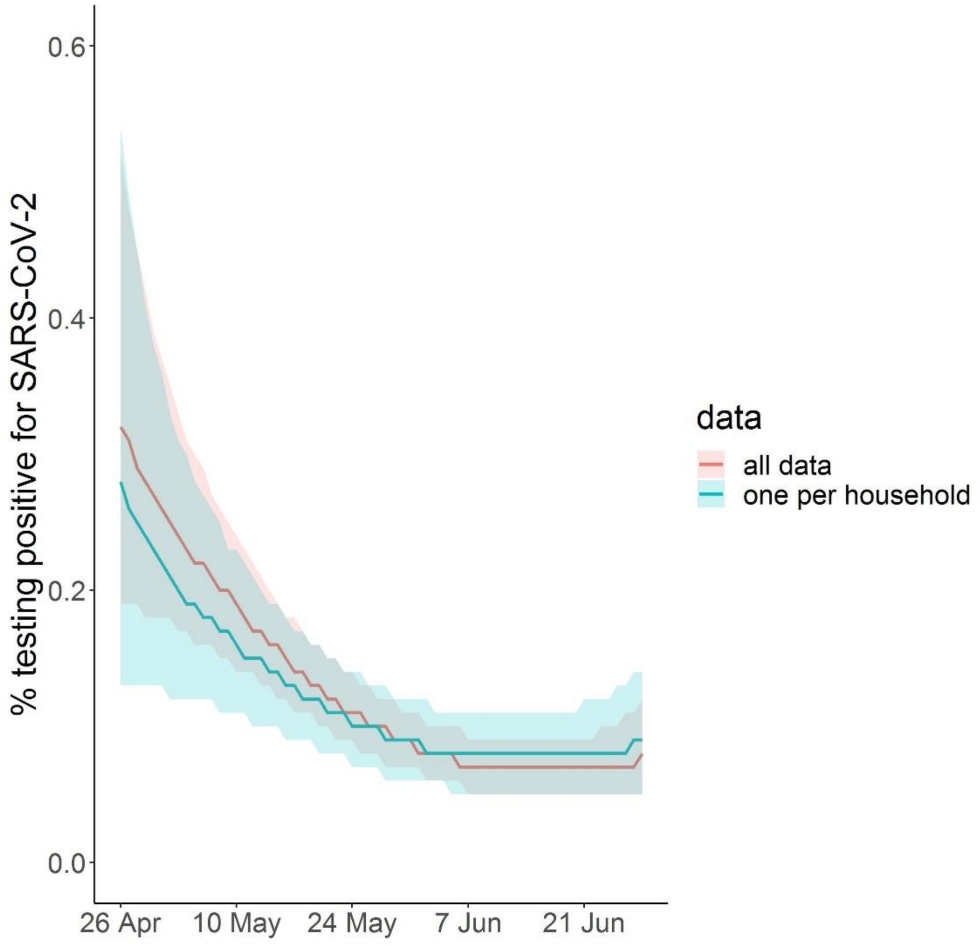


**Figure S1. Comparison of using all data (34,992 participants) for the multi-level regression with post-stratification for estimation of the percentage of inhabitants testing positive over time versus use one randomly selected person per household (16,772 participants).** The shaded area falls within the 95% credible intervals.

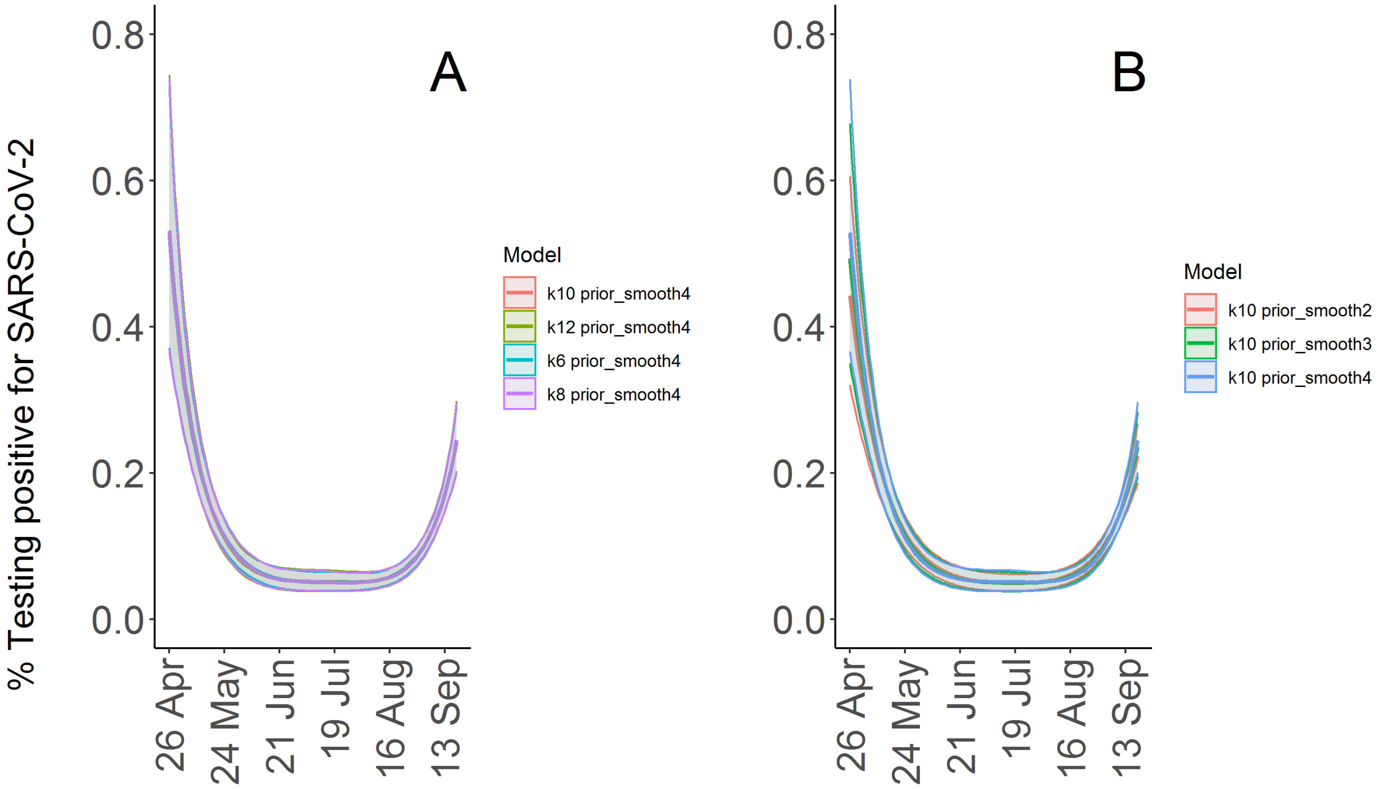


**Figure S2.** Comparison of models using different values for k (6, 8, 10, and 12, higher values resulted in divergent transitions) (A) and for the prior of the standard deviation of the smooth (normal prior with location 2, 3, and 4) for estimating the percentage of the population living in private households testing positive for SARS-CoV-2 with and without reporting symptoms.

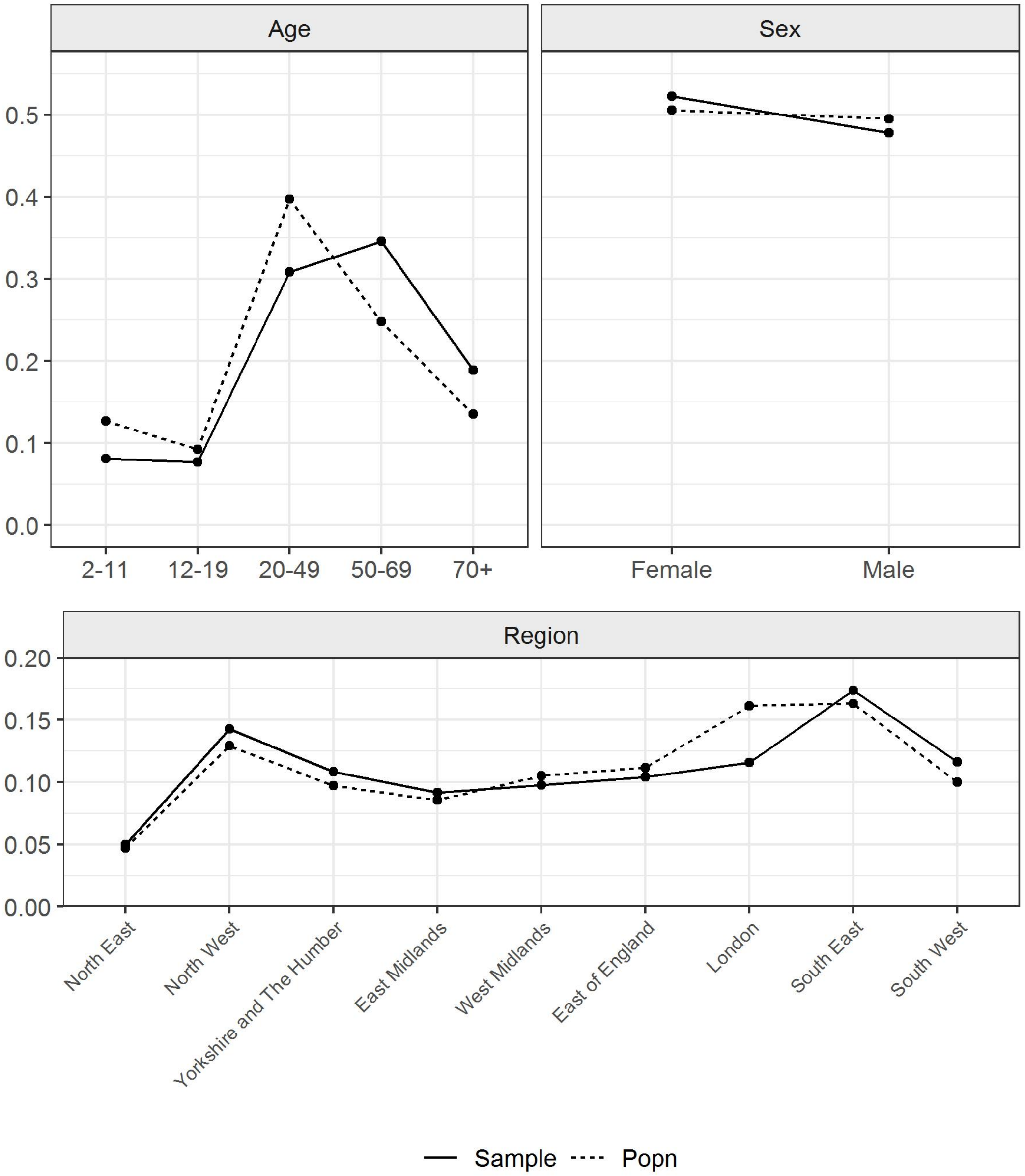


**Figure S3.** Representativeness of sample in terms of age and sex. Proportion of sample (solid line) and population in England (dashed line) within age- and sex-categories and regions.

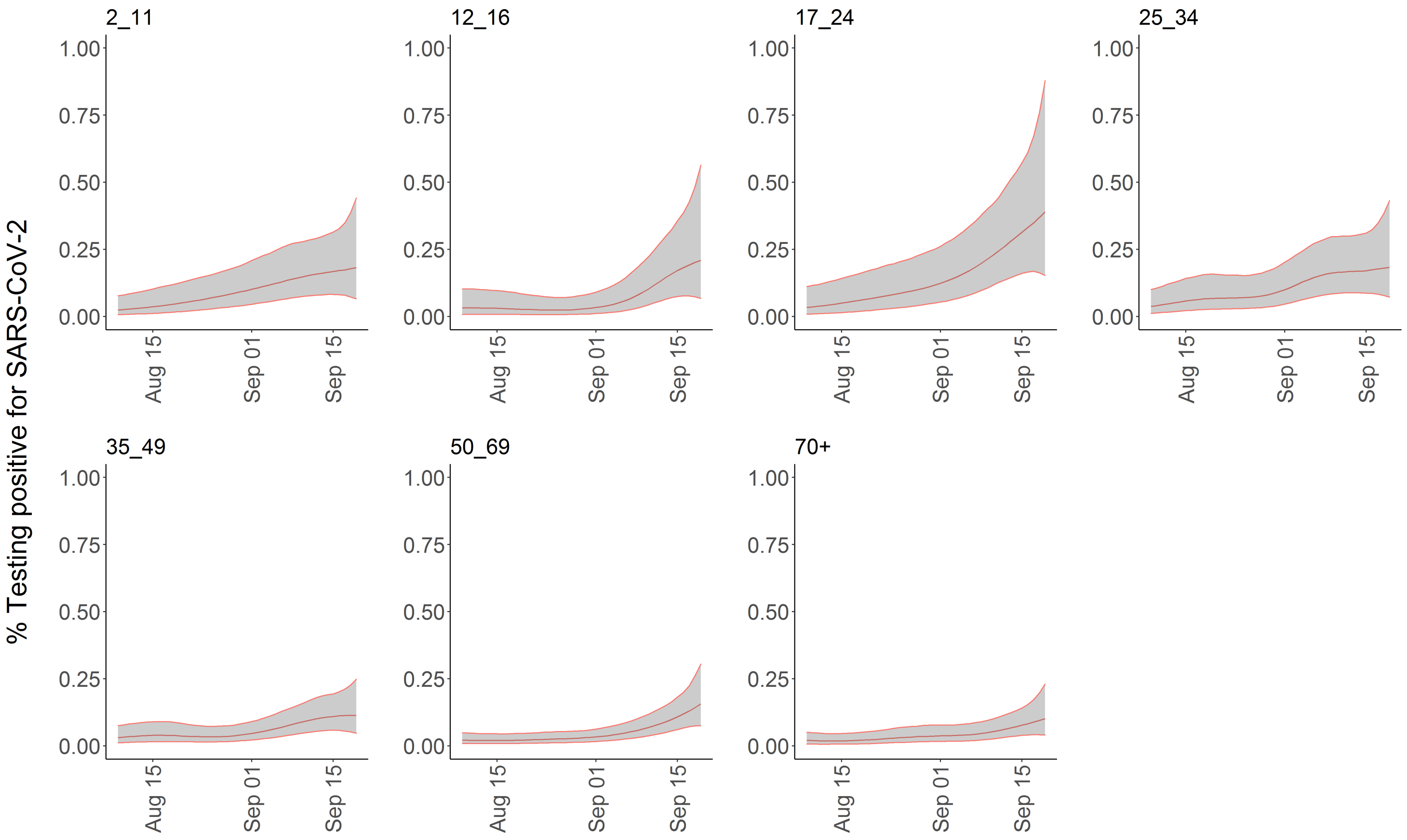


**Figure S4. Percentage of population within age (in years) subgroups testing positive for SARS-CoV-2.** Estimates are from a multilevel Bayesian GAM without post-stratification. The shaded area falls within the 95% credible intervals.

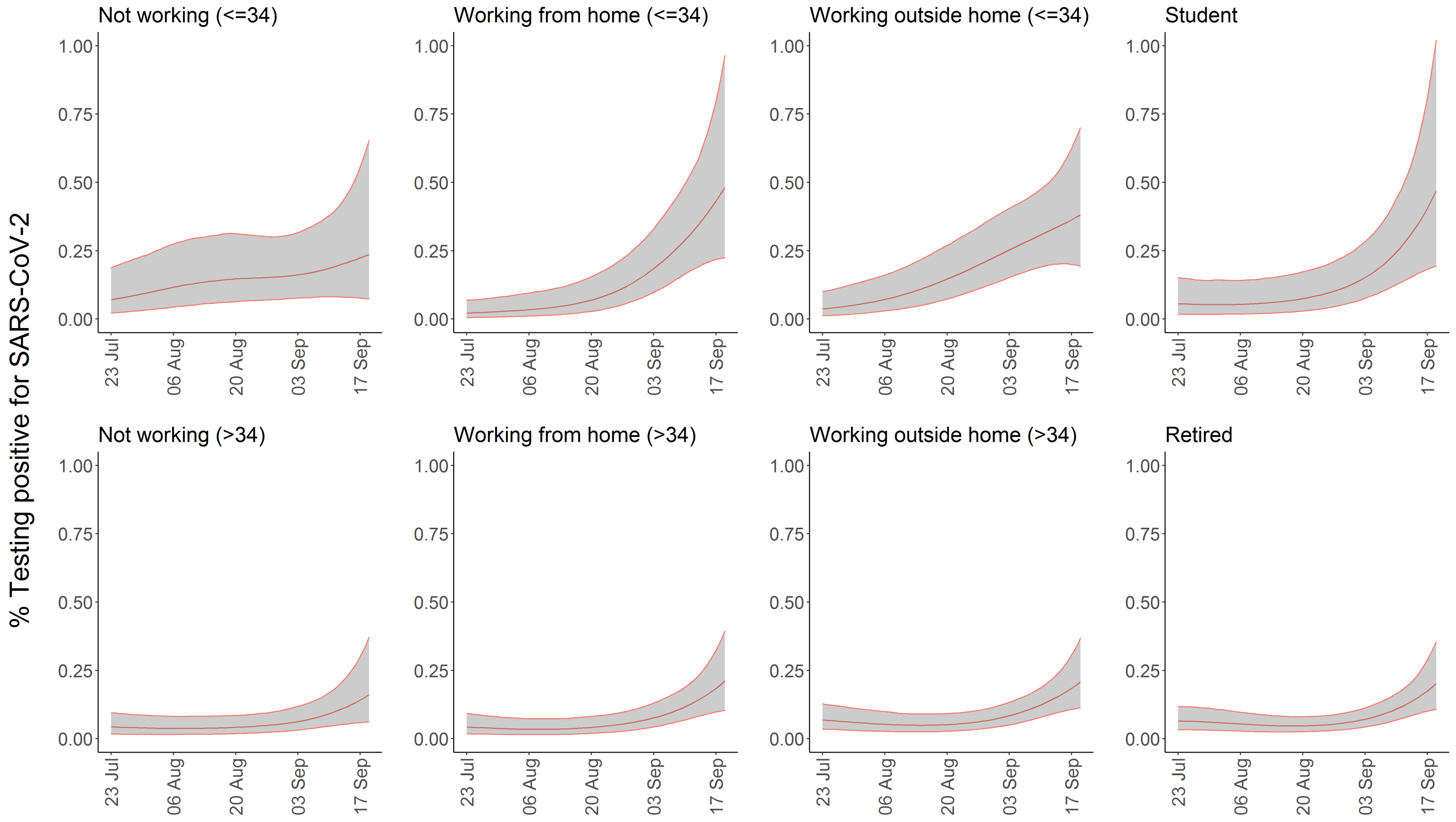


**Figure S5**. Percentage of population testing positive for SARS-CoV-2 by work location. Estimates are from a multilevel Bayesian GAM without post-stratification. The shaded area falls within the 95% credible intervals.

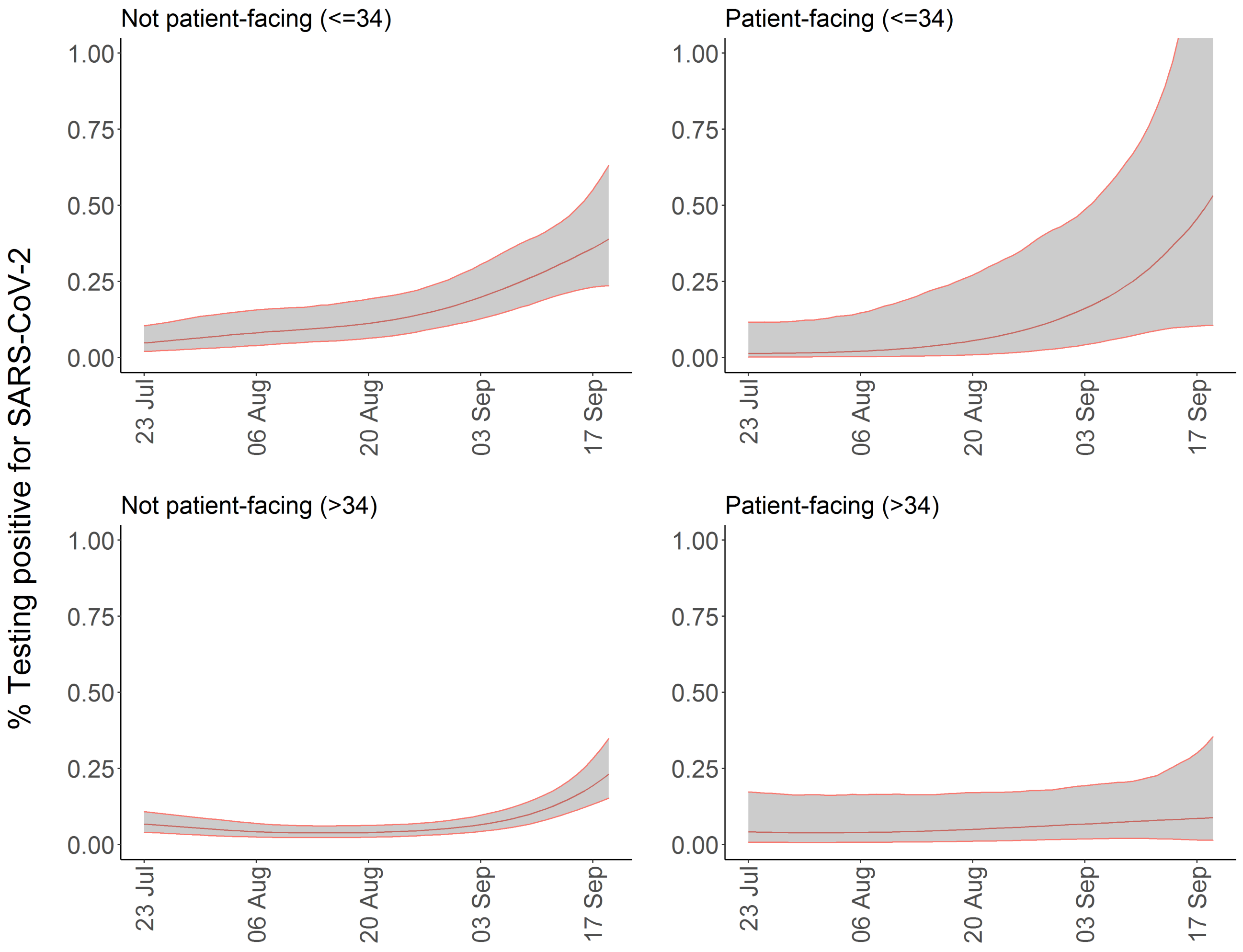


**Figure S6.** Percentage of population testing positive for SARS-CoV-2 by having a patient-facing role or not. Estimates are from a multilevel Bayesian GAM without post-stratification. The shaded area falls within the 95% credible intervals.

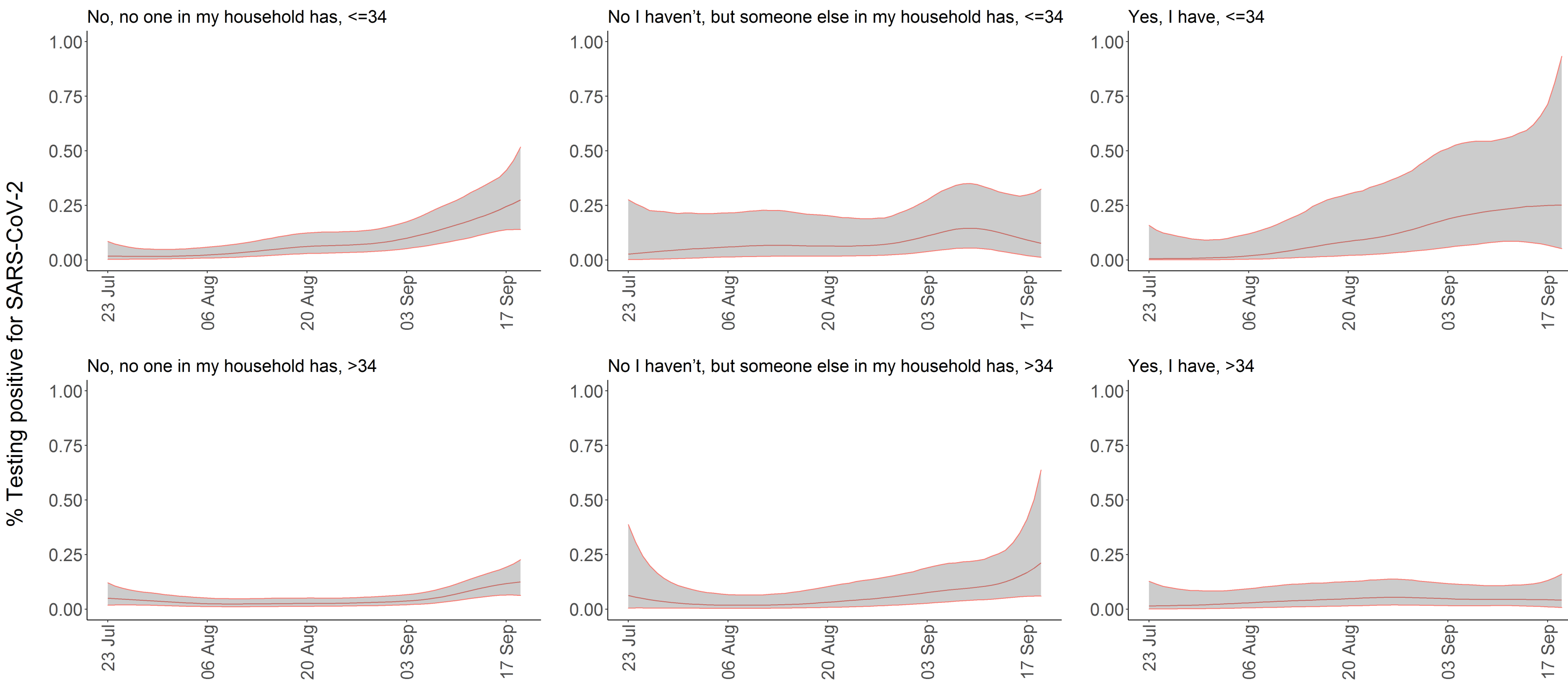


**Figure S7.** Percentage of population testing positive for SARS-CoV-2 by contact with hospital. Estimates are from a multilevel Bayesian GAM without post-stratification. The shaded area falls within the 95% credible intervals.

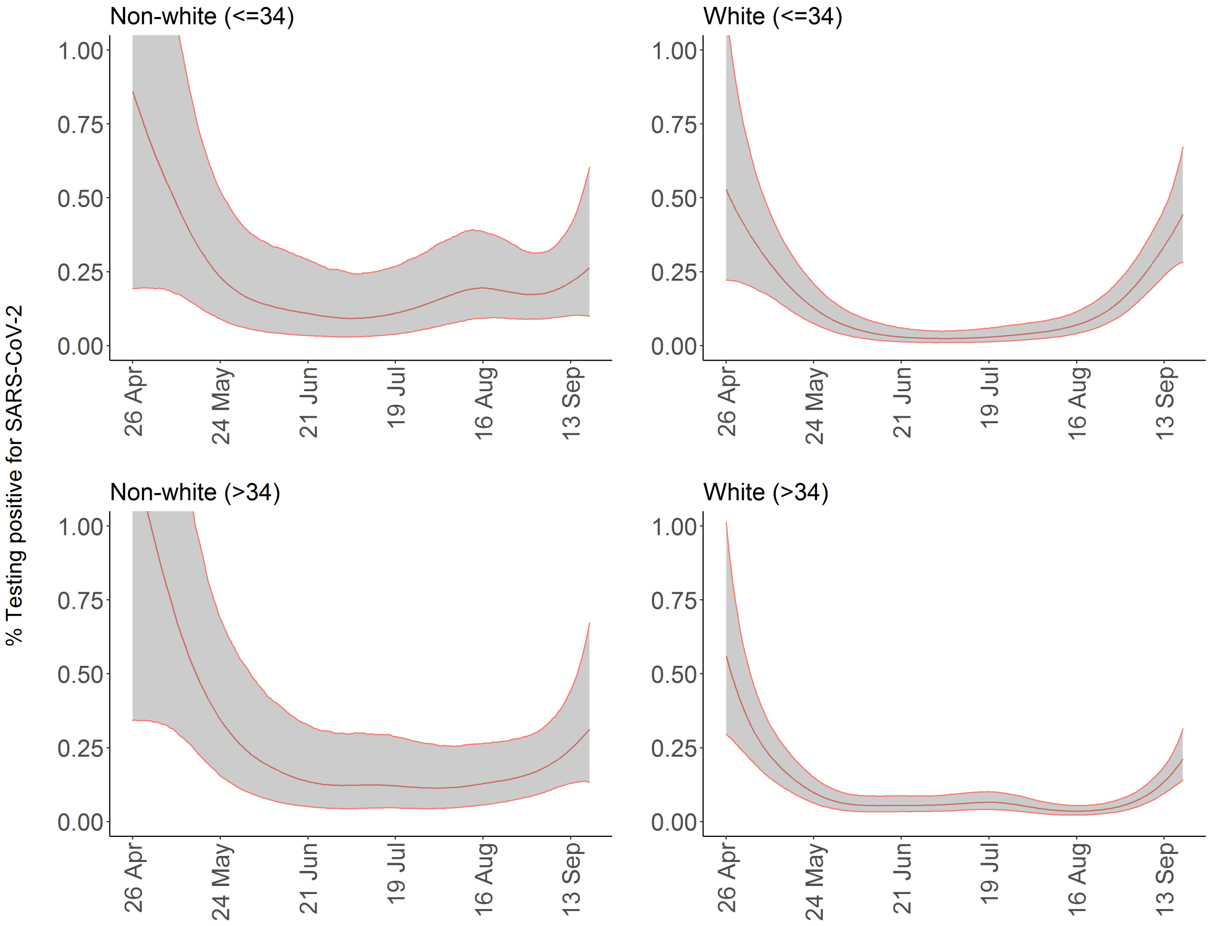


**Figure S8.** Percentage of population testing positive for SARS-CoV-2 by ethnicity (white / non-white). Estimates are from a multilevel Bayesian GAM without post-stratification. The shaded area falls within the 95% credible intervals.

### 7 References


1. https://cran.r-project.org/web/packages/rstanarm/vignettes/mrp.html
2. http://www.stat.columbia.edu/~gelman/research/unpublished/MRT(1).pdf
3. https://www.jstatsoft.org/article/view/v080i01

#### 8 Rstanarm code used for the regression model results from table S1 in the main paper and full details on all models considered

Model 1: Analysis from Table S1

Model 2: Model 1 + random intercept for household.

Model 3: Model 1 + hospital/care home contact added to the model. The relative exposure for contact with hospital and contact with care home reported in Table 1 come from this model. As these questions were only added since 8 May 2020, this regression is restricted to subset of data starting at 8 May.

Model 4: A reduced model with less covariates than model 1.

| | Model 1: Analysis from main text (table 1) | Model 2: model 1 + random intercept for household | Model 3: model 1 + hospital/carehome contact | Model 4: reduced model |
|---|---|---|---|---|
| *priors for covariables* | norm(0,1) | norm(0,1) | norm(0,1) | norm(0,1) |
| *prior for intercept* | norm(0,10) | norm(0,10) | norm(0,10) | norm(0,10) |
| *prior for covariance matrix (decov function in rstanarm)* | gamma(1,1) | gamma(1,1) | gamma(1,1) | gamma(1,1) |
| *regression formula* | stan_gamm4(result ~sex + s(study_day, by=region, k=5) + s(age, k=5) + work_location + patient_facing + resident_facing + ethnicity + householdsize + numchild, random= ~ (1 \| region), family=binomial(link="cloglog"), data=adults, iter=3000, cores=4, prior=normal(0,1), control=list(adapt_delta=0.95)) | stan_gamm4(result ~sex + s(study_day, by=region, k=5) + s(age, k=5) + work_location + patient_facing + resident_facing + ethnicity + householdsize + numchild, random= ~ (1 \| region/household_id), family=binomial(link="cloglog"), data=adults, iter=3000, cores=4, prior=normal(0,1), control=list(adapt_delta=0.95)) | stan_gamm4(result ~sex + s(study_day, by=region, k=5) + s(age, k=5) + work_location + patient_facing + resident_facing + ethnicity + householdsize + numchild + contact_hospital + contact_carehome, random= ~ (1 \| region), family=binomial(link="cloglog"), data=adults, iter=3000, cores=4, prior=normal(0,1), control=list(adapt_delta=0.95)) | stan_gamm4(result ~sex + s(study_day, by=region, k=5) + s(age, k=5) + work_location + patient_facing + resident_facing, random= ~ (1 \| region), family=binomial(link="cloglog"), data=adults, iter=3000, cores=4, prior=normal(0,1), control=list(adapt_delta=0.95)) |
| | Relative exposure (95% CrI) | Relative exposure (95% CrI) | Relative exposure (95% CrI) | Relative exposure (95% CrI) |
| Intercept | 0.00045 (0.0002 to 0.00092) | 0.00002 (0.000004 to 0.00005) | 0.0005 (0.002 to 0.0011) | 0.00043 (0.0002 to 0.00079) |
| Female | 0.84 (0.57 to 1.25) | 0.89 (0.59 to 1.35) | 0.77 (0.49 to 1.19) | 0.82(0.55 to 1.22) |
| *work location* | | | | |
| Working outside of your home | 2.47 (1.40 to 4.55) | 2.34 (1.24 to 4.51) | 2.11 (1.09 to 4.28) | 2.73 (1.55 to 4.96) |
| Both (working from home and working outside of your home) | 1.43 (0.53 - 3.54) | 1.54 (0.55 to 4.05) | 0.88 (0.24 to 2.60) | 1.52 (0.54 to 3.76) |
| Not applicable | 2.09 (1.20 to 3.75) | 2.07 (1.14 to 4.03) | 2.16 (1.17 to 4.11) | 2.08 (1.18 to 3.78) |
| *Patient/resident facing* | | | | |
| Patient-facing | 4.06 (2.37 to 6.72) | 4.73 (2.38 to 9.29) | 3.76 (1.92 to 7.02) | 4.24 (2.47 to 6.99) |
| Resident-facing | 2.35 (0.85 to 5.27) | 1.79 (0.57 to 5.06) | 0.91 (0.19 to 3.17) | 2.39 (0.88 to 5.52) |
| *Ethnicity* | | | | |
| Asian | 1.89 (0.87 to 3.64) | 1.56 (0.50 to 4.47) | 1.45 (0.56 to 3.30) | |
| Black | 1.04 (0.28 to 3.07) | 1.38 (0.32 to 5.46) | 1.30 (0.31 to 3.97) | |
| Mixed | 0.46 (0.09 to 1.84) | 0.54 (0.09 to 2.46) | 0.49 (0.09 to 1.98) | |
| Other | 7.5 (2.86 to 16.50) | 3.41 (0.74 to 13.09) | 6.59 (2.14 to 16.07) | |
| *Household size* | | | | |
| 2 | 0.62 (0.35 to 1.09) | 0.64 (0.32 to 1.30) | 0.47 (0.24 to 0.91) | |
| 3 | 1.51 (0.83 to 2.73) | 1.28 (0.59 to 2.88) | 1.61 (0.83 to 3.11) | |
| 4 | 1.36 (0.68 to 2.63) | 1.38 (0.59 to 3.36) | 1.58 (0.76 to 3.21) | |
| 5 or more | 1.25 (0.51 to 2.88) | 1.39 (0.47 to 4.10) | 1.36 (0.49 to 3.52) | |
| *Number of children in household* | | | | |
| 1 | 1.01 (0.59 to 1.73) | 1.12 (0.52 to 2.39) | 0.80 (0.43 to 1.46) | |
| 2 | 0.44 (0.20 to 0.94) | 0.52 (0.19 to 1.38) | 0.40 (0.16 to 0.89) | |
| 3 or more | 1.65 (0.64 to 4.17) | 1.61 (0.46 to 5.19) | 1.38 (0.47 to 3.83) | |
| *Contact with hospital* | | | | |
| Yes, I have | | | 2.18 (1.09 to 4.18) | |
| No I haven't, but someone else in my household has | | | 1.99 (0.86 to 4.13) | |
| *Contact with care home* | | | | |
| Yes, I have | | | 0.77 (0.18 to 2.79) | |
| No I haven't, but someone else in my household has | | | 0.48 (0.10 to 1.88) | |

#### 8 STROBE checklist

STROBE Statement—checklist of items that should be included in reports of observational studies

| | Item No. | Recommendation | Page No. |
|---|---|---|---|
| **Title and abstract** | 1 | (*a*) Indicate the study's design with a commonly used term in the title or the abstract | 1 |
| | | (*b*) Provide in the abstract an informative and balanced summary of what was done and what was found | 2 |
| **Introduction** | | | |
| Background/rationale | 2 | Explain the scientific background and rationale for the investigation being reported | 4 |
| Objectives | 3 | State specific objectives, including any prespecified hypotheses | 4 |
| **Methods** | | | |
| Study design | 4 | Present key elements of study design early in the paper | 4-5 |
| Setting | 5 | Describe the setting, locations, and relevant dates, including periods of recruitment, exposure, follow-up, and data collection | 4-5 |
| Participants | 6 | (*a*) *Cohort study*—Give the eligibility criteria, and the sources and methods of selection of participants. Describe methods of follow-up<br>*Case-control study*—Give the eligibility criteria, and the sources and methods of case ascertainment and control selection. Give the rationale for the choice of cases and controls<br>*Cross-sectional study*—Give the eligibility criteria, and the sources and methods of selection of participants | 4-5 |
| | | (*b*) *Cohort study*—For matched studies, give matching criteria and number of exposed and unexposed<br>*Case-control study*—For matched studies, give matching criteria and the number of controls per case | |
| Variables | 7 | Clearly define all outcomes, exposures, predictors, potential confounders, and effect modifiers. Give diagnostic criteria, if applicable | 5-7 |
| Data sources/ measurement | 8* | For each variable of interest, give sources of data and details of methods of assessment (measurement). Describe comparability of assessment methods if there is more than one group | 4-7 |
| Bias | 9 | Describe any efforts to address potential sources of bias | 6-7 |
| Study size | 10 | Explain how the study size was arrived at | 4-5 |

Continued on next page

| | | | |
|---|---|---|---|
| Quantitative variables | 11 | Explain how quantitative variables were handled in the analyses. If applicable, describe which groupings were chosen and why | 5-6 |
| Statistical methods | 12 | (*a*) Describe all statistical methods, including those used to control for confounding | 5-7 |
| | | (*b*) Describe any methods used to examine subgroups and interactions | 6-7 |
| | | (*c*) Explain how missing data were addressed | 5-6 |
| | | (*d*) *Cohort study*—If applicable, explain how loss to follow-up was addressed<br>*Case-control study*—If applicable, explain how matching of cases and controls was addressed<br>*Cross-sectional study*—If applicable, describe analytical methods taking account of sampling strategy | NA |
| | | (*e*) Describe any sensitivity analyses | 6-7 |
| **Results** | | | |
| Participants | 13* | (a) Report numbers of individuals at each stage of study—eg numbers potentially eligible, examined for eligibility, confirmed eligible, included in the study, completing follow-up, and analysed | 7 |
| | | (b) Give reasons for non-participation at each stage | NA |
| | | (c) Consider use of a flow diagram | NA |
| Descriptive data | 14* | (a) Give characteristics of study participants (eg demographic, clinical, social) and information on exposures and potential confounders | Figure 1, Table 1 |
| | | (b) Indicate number of participants with missing data for each variable of interest | 5-6 |
| | | (c) *Cohort study*—Summarise follow-up time (eg, average and total amount) | 7 |
| Outcome data | 15* | *Cohort study*—Report numbers of outcome events or summary measures over time | 7, Figure 2 |
| | | *Case-control study*—Report numbers in each exposure category, or summary measures of exposure | |
| | | *Cross-sectional study*—Report numbers of outcome events or summary measures | *7* |
| Main results | 16 | (*a*) Give unadjusted estimates and, if applicable, confounder-adjusted estimates and their precision (eg, 95% confidence interval). Make clear which confounders were adjusted for and why they were included | 7-8, Table 1 |
| | | (*b*) Report category boundaries when continuous variables were categorized | Table 1 |
| | | (*c*) If relevant, consider translating estimates of relative risk into absolute risk for a meaningful time period | NA |

Continued on next page

| | | | |
|---|---|---|---|
| Other analyses | 17 | Report other analyses done—eg analyses of subgroups and interactions, and sensitivity analyses | 7-8 |
| **Discussion** | | | |
| Key results | 18 | Summarise key results with reference to study objectives | 9 |
| Limitations | 19 | Discuss limitations of the study, taking into account sources of potential bias or imprecision. Discuss both direction and magnitude of any potential bias | 10-11 |
| Interpretation | 20 | Give a cautious overall interpretation of results considering objectives, limitations, multiplicity of analyses, results from similar studies, and other relevant evidence | 11 |
| Generalisability | 21 | Discuss the generalisability (external validity) of the study results | 9-11 |
| **Other information** | | | |
| Funding | 22 | Give the source of funding and the role of the funders for the present study and, if applicable, for the original study on which the present article is based | |

*Give information separately for cases and controls in case-control studies and, if applicable, for exposed and unexposed groups in cohort and cross-sectional studies.

**Note:** An Explanation and Elaboration article discusses each checklist item and gives methodological background and published examples of transparent reporting. The STROBE checklist is best used in conjunction with this article (freely available on the Web sites of PLoS Medicine at http://www.plosmedicine.org/, Annals of Internal Medicine at http://www.annals.org/, and Epidemiology at http://www.epidem.com/). Information on the STROBE Initiative is available at www.strobe-statement.org.
